# The impact of the SYNTAX score on the incidence of in-hospital cardiac arrest in patients with acute coronary syndrome undergoing percutaneous coronary intervention: a retrospective cohort study

**DOI:** 10.64898/2026.09.23.26363831

**Authors:** Kui Li, Hongxu Chen, Haowen Ye, Jun Guo

**Affiliations:** Department of Cardiology, The First Hospital of Jinan University, Guangzhou, China; The First Clinical Medical College, Jinan University, Guangzhou, China

**Keywords:** SYNTAX score, in-hospital cardiac arrest, acute coronary syndrome, retrospective cohort study, percutaneous coronary intervention

## Abstract

**Background:** The SYNTAX score (SS) quantifies the severity and complexity of coronary artery lesions and serves as an independent predictor of major adverse cardiovascular events and mortality in patients with acute coronary syndrome (ACS). However, few studies have examined the association between SS and in-hospital cardiac arrest (IHCA) in this population. Therefore, this study aims to elucidate the relationship between SS and the incidence of IHCA in patients with ACS.

**Methods:** This study consecutively enrolled 3,224 patients undergoing percutaneous coronary intervention (PCI). Logistic regression and restricted cubic spline (RCS) analyses were employed to examine the association between SS and the incidence of IHCA. Participants were stratified into three groups based on SS: Group T1 (SS ≤ 10, N = 1,097), Group T2 (10 < SS ≤ 18, N = 1,089), and Group T3 (SS > 18, N = 1,038). The primary endpoint was the incidence of IHCA.

**Results:** The overall incidence of IHCA was 4% (N=147). After adjusting for covariates, SS was significantly associated with the incidence of IHCA in ACS patients treated with PCI (OR = 1.0400; 95% confidence interval (CI) = 1.0200–1.0700; *p* < 0.001). Compared with the T1 group, the T3 group exhibited an increased risk of IHCA (OR = 2.1800; 95% CI = 1.2900–3.6900; *p* = 0.0040). Subgroup analyses revealed that, after covariate adjustment, patients with ST-segment elevation myocardial infarction (STEMI) had an increased risk of IHCA (OR = 1.0700; 95% CI = 1.0300–1.1100; *p* < 0.001). Similarly, both patients with diabetes mellitus (DM) (OR = 1.0400; 95% CI = 1.000–1.0700; *p* = 0.0270) and those without DM (OR = 1.0600; 95% CI = 1.0200–1.1000; *p* = 0.0020) showed an increased risk. Furthermore, elderly patients (≥60 years) (OR = 1.0300; 95% CI = 1.0100–1.0700; *p* = 0.0210) and non-elderly patients (<60 years) (OR = 1.0500; 95% CI = 1.000–1.1000; *p* = 0.0470) had an increased risk, as did male patients (OR = 1.0500; 95% CI = 1.0200–1.0700; *p* = 0.0020). RCS analysis indicated a dose-response relationship between IHCA incidence and SS (nonlinear *p*-value = 0.906), with the risk of IHCA increasing when SS exceeded 23.975. Incorporating SS into the baseline risk model improved the predictive value for IHCA incidence in ACS patients undergoing PCI (Net reclassification improvement [NRI]: 0.3537 [0.1906–0.5167], *p* < 0.001; Integrated discriminant improvement [IDI]: 0.0104 [0.0025–0.0183], *p* = 0.0097).

**Conclusions:** Among patients with ACS undergoing PCI, SS is significantly associated with the incidence of IHCA. Consequently, SS may serve as an effective predictor of IHCA in this population.

## Background

ACS, encompassing unstable angina (UA), non-ST-segment elevation myocardial infarction (NSTEMI), and STEMI, is a leading cause of mortality^1,2^. Although PCI has significantly improved the overall prognosis of patients with ACS^3^, cardiac arrest remains the most lethal complication during hospitalization. Studies indicate that the incidence of IHCA among patients with ACS ranges from 1.50% to 6.2%; once it occurs, the in-hospital mortality rate can exceed 55%^4,5^. Advancing age, elevated blood glucose levels, and a heavier burden of cardiovascular disease are closely associated with the occurrence of IHCA and poor outcomes in patients with ACS^4–7^. Therefore, identifying patients at high risk for IHCA and refining risk stratification are of significant clinical importance. However, while tools such as the GRACE score are widely used in clinical practice to predict the risk of in-hospital mortality^8,9^, they do not adequately capture coronary anatomical features and thus fall short of fully meeting the clinical needs for IHCA risk stratification. In this context, SS can quantify the severity and complexity of coronary artery lesions and independently predict the risk of major adverse cardiovascular events and death in patients with ACS^10–12^. Notably, among elderly patients with ACS, the association between elevated SS levels and an increased risk of short-term mortality is particularly evident^13–15^. Previous studies have largely focused on the predictive value of SS for major adverse cardiovascular events or all-cause mortality^10,12,16^, whereas studies specifically examining the association between SS and IHCA in patients with ACS undergoing PCI remain scarce. Therefore, this study aims to clarify the relationship between SS and the incidence of IHCA in patients with ACS undergoing PCI.

## Methods

### Study design and population

This retrospective cohort study adhered to the Declaration of Helsinki and received approval from the hospital ethics committee, which waived the requirement for informed consent due to the retrospective nature of the research. Patients who underwent PCI between May 1, 2021, and May 1, 2025, were consecutively enrolled. Inclusion criteria comprised patients with ACS undergoing PCI. Patients with missing data were excluded (Figure 1). Ultimately, 3,224 ACS patients who underwent PCI were included in the final analysis. Participants were stratified into three groups based on the SS: Group T1 (SS ≤ 10, N = 1,097), Group T2 (10 < SS ≤ 18, N = 1,089), and Group T3 (SS > 18, N = 1,038). The primary endpoint was the incidence of IHCA.

**Figure 1.**
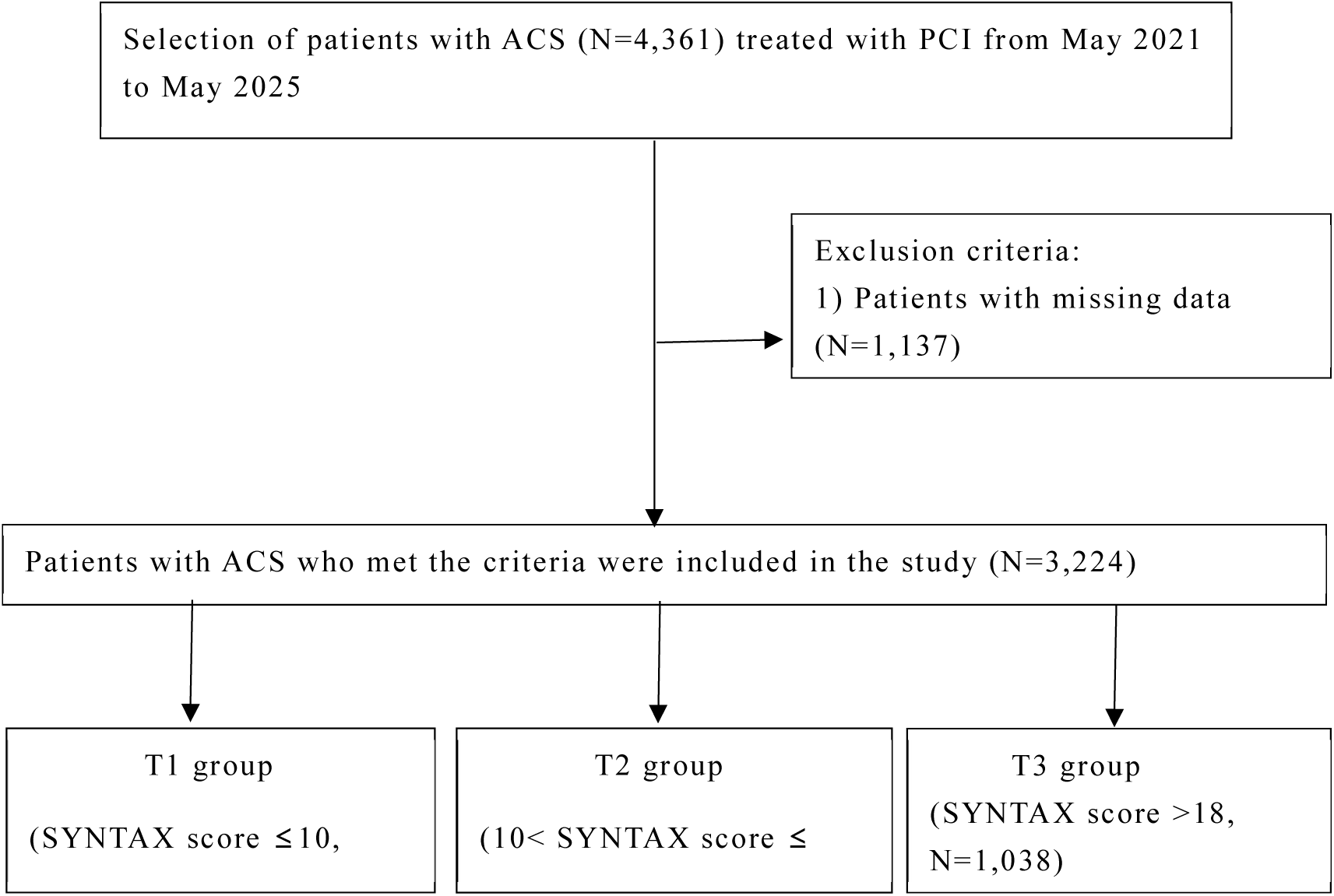
Patient inclusion flowchart.

### Data measurement and definitions

Baseline demographic and clinical data were retrospectively extracted from hospital medical records. Demographic variables included age, sex, body mass index (BMI), smoking status, comorbidities (hypertension, dyslipidemia, DM, history of stroke, family history, previous myocardial infarction (MI), previous PCI, and previous coronary artery bypass grafting (CABG). Clinical data encompassed admission systolic (SBP) and diastolic blood pressure (DBP); primary diagnosis (STEMI, NSTEMI, UA); imaging and procedural details (transradial approach, left main (LM) disease, left anterior descending (LAD) artery disease, left circumflex (LCX) artery disease, right coronary artery (RCA) disease, chronic total occlusion (CTO), number of diseased vessels, bifurcation lesions, number of stents, stent length, stent diameter, thrombolytic therapy, and rotational atherectomy); and laboratory results (left ventricular ejection fraction (LVEF), triglycerides (TG), total cholesterol (TC), high-density lipoprotein cholesterol (HDL-C), low-density lipoprotein cholesterol (LDL-C), admission blood glucose (ABG), glycated hemoglobin (HbA1c), hemoglobin (Hb), creatinine (Cr), uric acid, and high-sensitivity C-reactive protein (hs-CRP)). Medication regimens during hospitalization included insulin, oral hypoglycemic agents, aspirin, P2Y12 inhibitors, statins, beta-blockers, and angiotensin-converting enzyme inhibitors (ACEI) or angiotensin II receptor blockers (ARB). IHCA was defined as the administration of external chest compressions and/or defibrillation to inpatients^17^, with data collected from medical records. ABG referred to the first random blood glucose measurement within 24 hours of admission. BMI was calculated as weight (kg) divided by height (m) squared. The estimated glomerular filtration rate (eGFR) was calculated using the MDRD formula^18^: for men, eGFR = 186 × Cr-1.154 × age-0.203; for women, eGFR = 186 × Cr-1.154 × age-0.203 × 0.742. The SS quantitatively assessed coronary artery disease (CAD) severity in ACS patients. Based on this score, CAD patients were classified as low risk (≤22), moderate risk (23–32), and high risk (≥33)^19^. CAD was defined as ≥50% luminal stenosis in at least one major coronary artery (LAD, LCX, or RCA). Left main coronary artery disease was defined as ≥50% stenosis in the left main coronary artery. DM was defined as a history of type 2 DM or an HbA1c level ≥6.5%, whereas non-DM was defined as an HbA1c level <6.5%^20^. Stroke was defined as a history of cerebral hemorrhage, ischemic stroke, or transient ischemic attack.

### Statistical analyses

Continuous variables following a normal distribution are presented as mean ± standard deviation, while non-normally distributed variables are presented as median (interquartile range). Categorical variables are expressed as counts (percentages). For normally distributed continuous variables with homoscedasticity, analysis of variance (ANOVA) was used to compare the three groups; for variables violating these assumptions, the Kruskal-Wallis rank-sum test was employed. The chi-square test was used for categorical variables. Logistic regression analysis was performed to calculate the odds ratio (OR) and 95% CI for the relationship between SS and IHCA incidence. Model 1 was unadjusted, while Model 2 was a multivariate model. Multicollinearity between SS and other covariates was assessed using the generalized variance inflation factor (GVIF); covariates with a GVIF(1/2Df) value ≥ 2 (where Df represents degrees of freedom) were considered to exhibit significant multicollinearity. Least absolute shrinkage and selection operator (LASSO) regression was then used to select covariates. The following covariates were adjusted for in the multivariate logistic regression analyses for the overall population: age, DBP, stent diameter, eGFR, ABG, hs-CRP, HbA1c, LVEF, male sex, smoking, previous MI, LCX disease, RCA disease, bifurcation lesion, CTO disease, thrombolytic therapy, hypertension, dyslipidemia, previous stroke, insulin use, oral hypoglycemic drugs, aspirin, and P2Y12 inhibitors. RCS analysis examined the relationship between SS and IHCA incidence. Given that the correlation between SS and IHCA incidence was approximately linear both below and above the SS value corresponding to an OR of 1, a linear model was used to calculate the OR for each standard deviation increase in SS. The predictive ability of the logistic regression model for IHCA incidence was quantified using the area under the receiver operating characteristic (ROC) curve (AUC), calculated via the C-statistic. The DeLong test was used to compare AUCs between models. Additionally, the NRI and IDI were calculated to evaluate the additional predictive value of SS beyond identified risk factors. Subgroup analyses were performed using logistic regression based on primary diagnosis at admission, diabetes status, sex, and age. A two-sided *p*-value < 0.05 was considered statistically significant. All statistical analyses were performed using R version 4.2.3.

## Results

### Baseline characteristics

This study included 3,224 patients with ACS who underwent PCI. The median age was 63 (54, 71) years. The cohort comprised 2,302 male patients (71.4%), 1,737 patients with DM (53.9%), 690 patients with STEMI (21.4%), 776 patients with NSTEMI (24.1%), and 1,758 patients with UA (54.5%). Significant differences were observed among the three groups regarding age, SBP, DBP, previous stroke, previous MI, previous PCI, diagnosis on admission, transradial approach, LM disease, LAD disease, RCA disease, CTO disease, number of diseased vessels, bifurcation lesion, number of stents, length of stents, diameter of stents, rotablator, LVEF, TG, TC, LDL-C, SS, Hb, Cr, eGFR, hs-CRP, insulin, and statins (all *p* < 0.05) (Table 1).

**Table 1.**
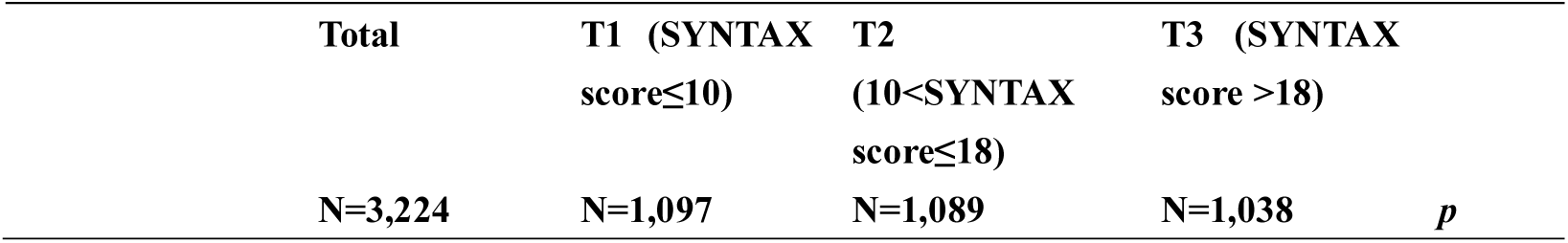
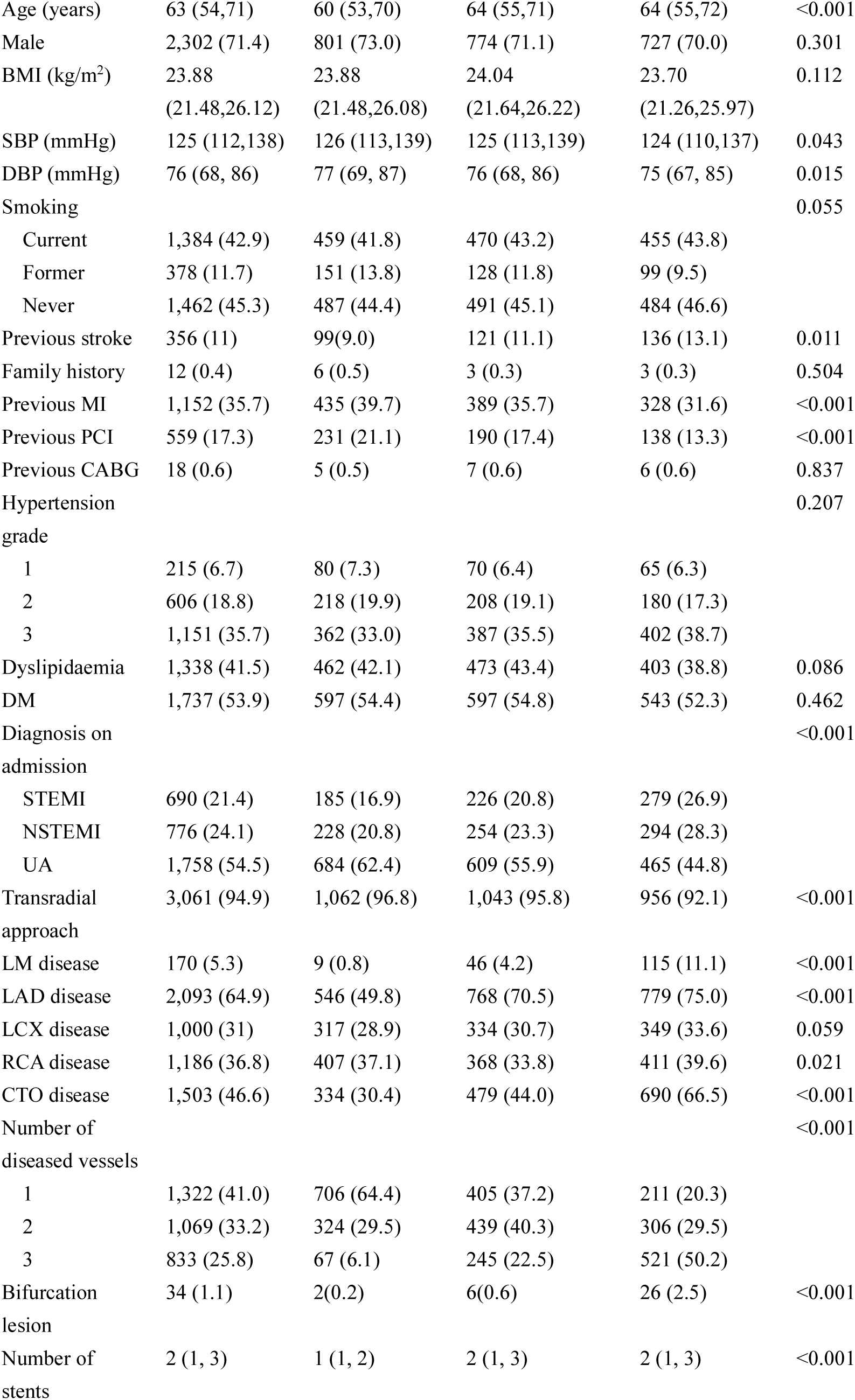
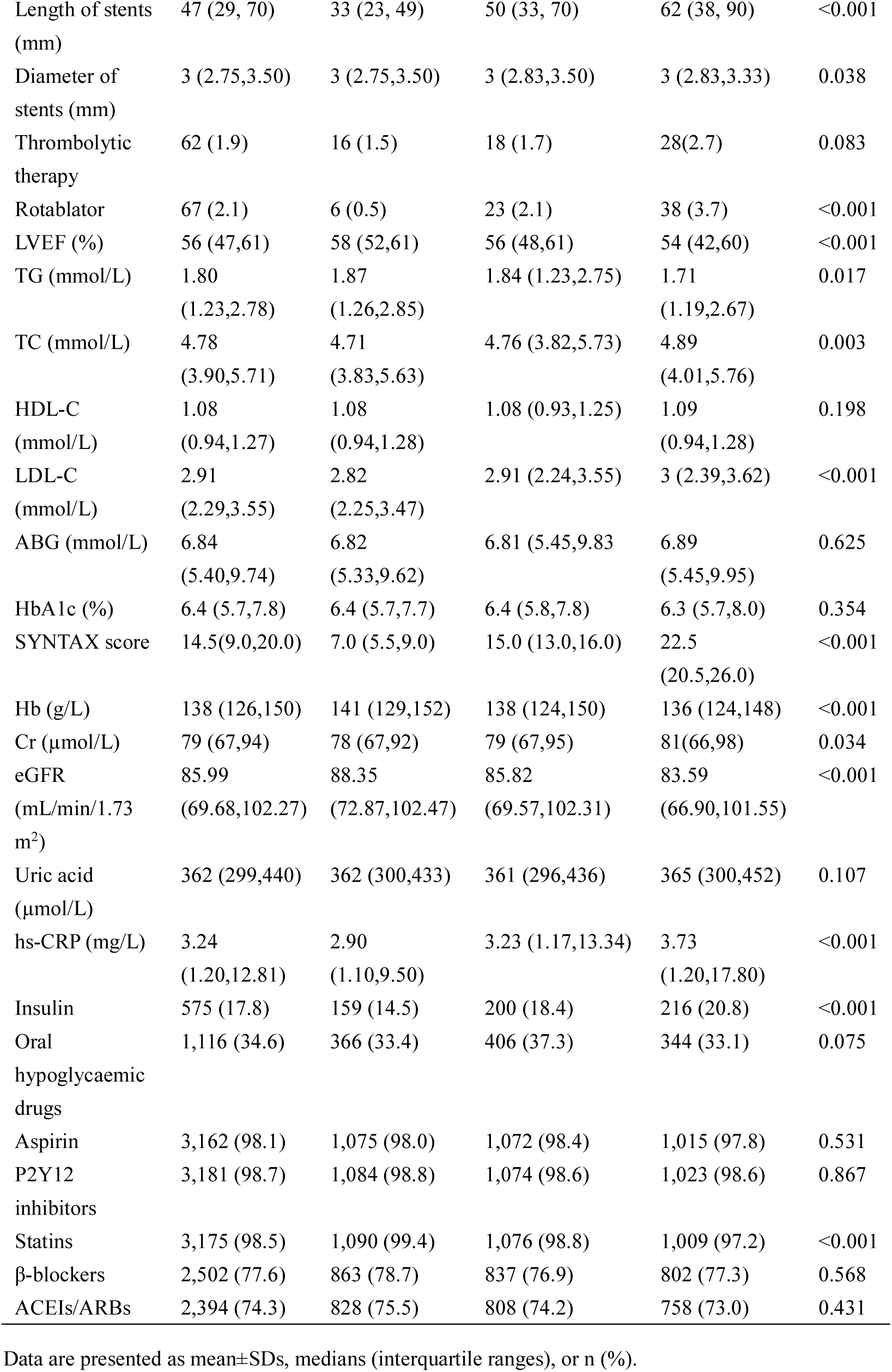
Demographic and clinical baseline data for the three groups.

### Clinical outcomes

The overall incidence of IHCA was 4.6% (N=147), with significant differences among the three groups (*p*<0.001) (Table 2). In Model 1, SS was significantly associated with IHCA incidence (OR = 1.0800; 95% CI = 1.0500–1.1000; *p* < 0.001) (Table 3). After adjusting for potential risk factors in Model 2, SS remained an independent risk factor for IHCA in patients with ACS (OR = 1.0400; 95% CI = 1.0200– 1.0700; *p* < 0.001). In Model 1, the IHCA incidence in the T3 group was 4.22 times that of the T1 group (OR = 4.2200; 95% CI = 2.6400–6.7400; *p* < 0.001). In Model 2, the incidence in the T3 group was 2.18 times that of the T1 group (OR = 2.1800; 95% CI = 1.2900–3.6900; *p* = 0.0040). RCS analysis indicated a dose-response relationship between SS and IHCA incidence, even after adjusting for confounders in Model 2 (nonlinear *p*-value = 0.906) (Figure 2). When SS < 23.975, the OR for IHCA incidence changed gradually. When SS > 23.975, the OR increased significantly. Specifically, when SS < 23.975, the OR per standard deviation (SD) for predicted IHCA incidence was 1.3 (1.0300–1.6300); when SS > 23.975, the OR per SD was 0.999 (0.6250–1.5100) (Table 4).

**Figure 2.**
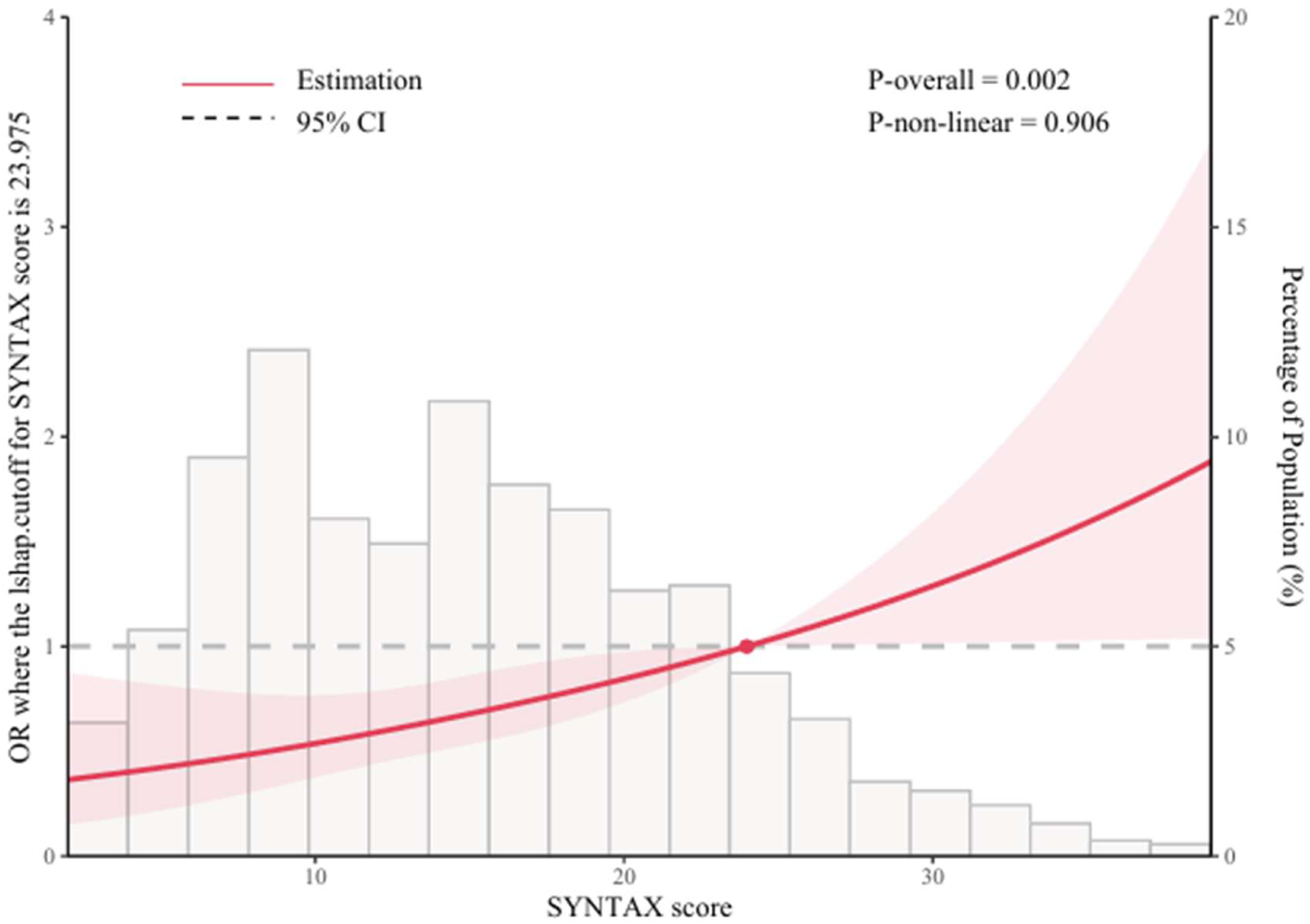
Relationship between SYNTAX score and incidence of in-hospital cardiac arrest in patients with ACS. Only 95% of the data is displayed. Odds ratios are indicated by solid lines and 95% CIs by shaded areas. acute coronary syndrome, ACS.

**Table 2.** Incidence of in-hospital cardiac arrest in the three groups.

| Endpoint | Total (N=3,224) | T1 (N=1,097) | T2 (N=1,089) | T3 (N=1,038) | *p* |
| --- | --- | --- | --- | --- | --- |
| IHCA | 147 (4.6%) | 23 (2.1%) | 38 (3.5%) | 86 (8.3%) | <0.001 |

In-Hospital Cardiac Arrest, IHCA

**Table 3.** Relationship between the SYNTAX score and incidence of in-hospital cardiac arrest.

|  | Events/N | Model 1 |  |  | Model 2 |  |  |
| --- | --- | --- | --- | --- | --- | --- | --- |
|  |  | OR | 95% CI | *p* | OR | 95% CI | *p* |
| SYNTAX score | 147/3,224 | 1.0800 | 1.0500-1.1000 | <0.001 | 1.0400 | 1.0200-1.0700 | <0.001 |
| T1 | 23/1,097 | Reference |  |  | Reference |  |  |
| T2 | 38/1,089 | 1.6900 | 1.000-2.8500 | 0.0500 | 1.3200 | 0.7400-2.3400 | 0.3430 |
| T3 | 86/1,038 | 4.2200 | 2.6400-6.7400 | <0.001 | 2.1800 | 1.2900-3.6900 | 0.0040 |
| *p* for trend |  |  |  | <0.001 |  |  | 0.0015 |

Odds Ratio, OR; Confidence Interval, CI; *p*-value, *p*

**Table 4.** Relationship between SYNTAX score (per 1 SD) and in-hospital cardiac arrest incidence in PCI-treated ACS patients.

|  | OR per SD | 95% CI |
| --- | --- | --- |
| SYNTAX score $\geq 23.975$ | 0.9990 | 0.6250-1.5100 |
| SYNTAX score $< 23.975$ | 1.3000 | 1.0300-1.6300 |

Odds Ratio, OR; Confidence Interval, CI; Standard Deviation, SD;

### Subgroup analyses

Subgroup analyses examined the association between SS and IHCA incidence across populations stratified by primary diagnosis at admission (STEMI, NSTEMI, and UA), diabetes status (DM and without DM), gender (male and female), and age (≥60 and <60).

Table 5 presents the relationship between SS and IHCA incidence in patients with STEMI, NSTEMI, and UA. SS did not interact with IHCA incidence across these subgroups (*p* = 0.4803). In the STEMI subgroup, Model 1 showed a significant association between SS and IHCA risk (OR = 1.0800; 95% CI = 1.0500–1.1100; *p* < 0.001), with the T3 group having 4.37 times the incidence of the T1 group (OR = 4.3700; 95% CI = 2.0100–9.4900; *p* < 0.001). Model 2 also demonstrated a significant association (OR = 1.0700; 95% CI = 1.0300–1.1100; *p* < 0.001), with the T3 group having 3.45 times the incidence of the T1 group (OR = 3.4500; 95% CI = 1.4800–8.0100; *p* = 0.0040).

**Table 5.**
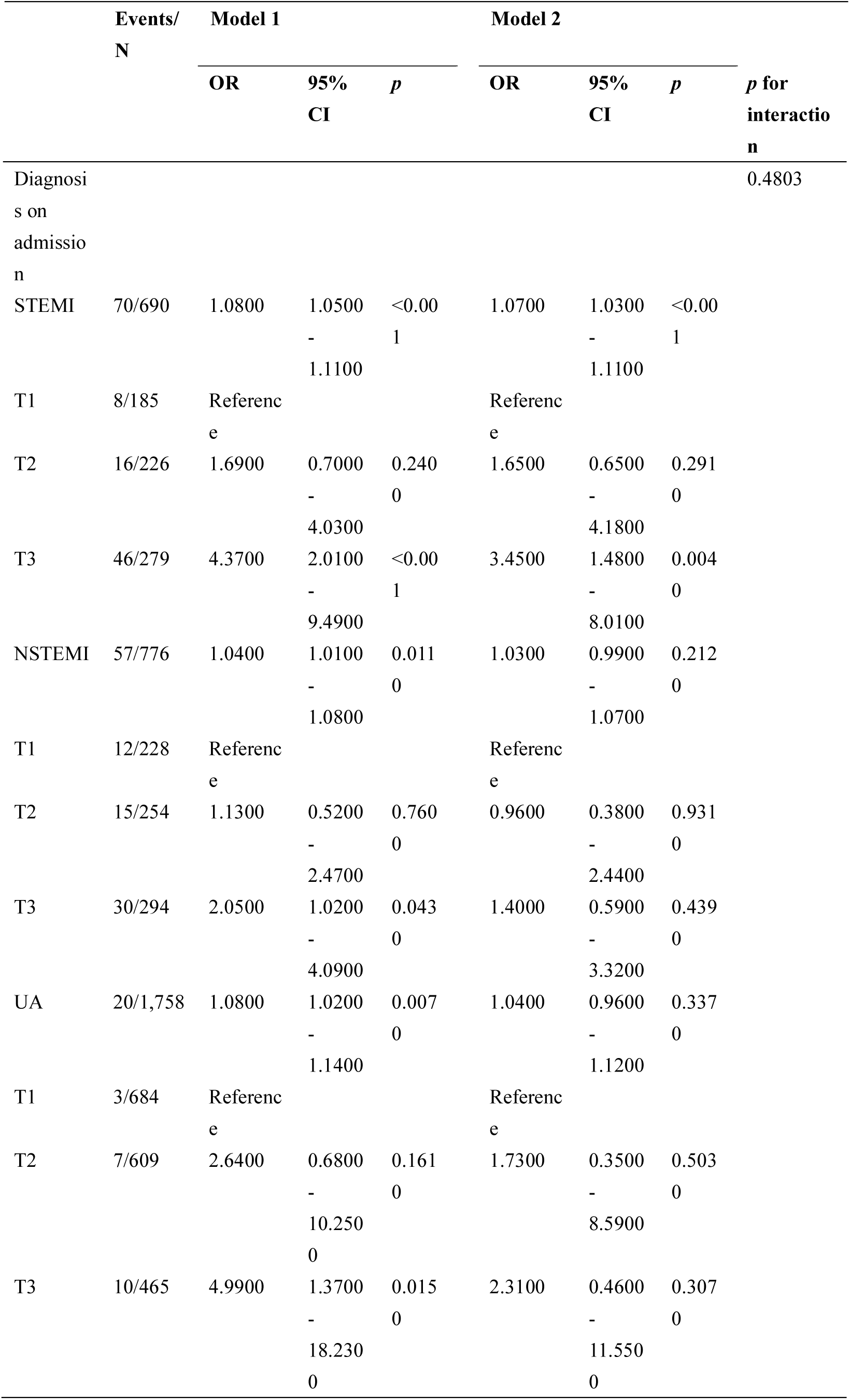
Relationship between SYNTAX score and in-hospital cardiac arrest incidence in patients with STEMI, NSTEMI, and UA.

Table 6 shows the relationship between SS and IHCA incidence in patients with and without diabetes. No interaction was observed between diabetes status and SS regarding IHCA incidence (*p*=0.3519). Among patients with DM, Model 1 indicated a significant association between SS and IHCA risk (OR = 1.0800; 95% CI = 1.0500–1.1100; *p* < 0.001), with the T3 group having 4.51 times the incidence of the T1 group (OR = 4.5100; 95% CI = 2.3600–8.6100; *p* < 0.001). Model 2 also showed a significant association (OR = 1.0400; 95% CI = 1.000–1.0700; *p* = 0.0270), with the T3 group having 2.16 times the incidence of the T1 group (OR = 2.1600; 95% CI = 1.0400–4.4900; *p* = 0.0390). Among patients without DM, both Model 1 (OR = 1.0700; 95% CI = 1.0400–1.1100; *p* < 0.001) and Model 2 (OR = 1.0600; 95% CI = 1.0200–1.1000; *p* = 0.0020) showed that SS was significantly associated with IHCA incidence.

**Table 6.**
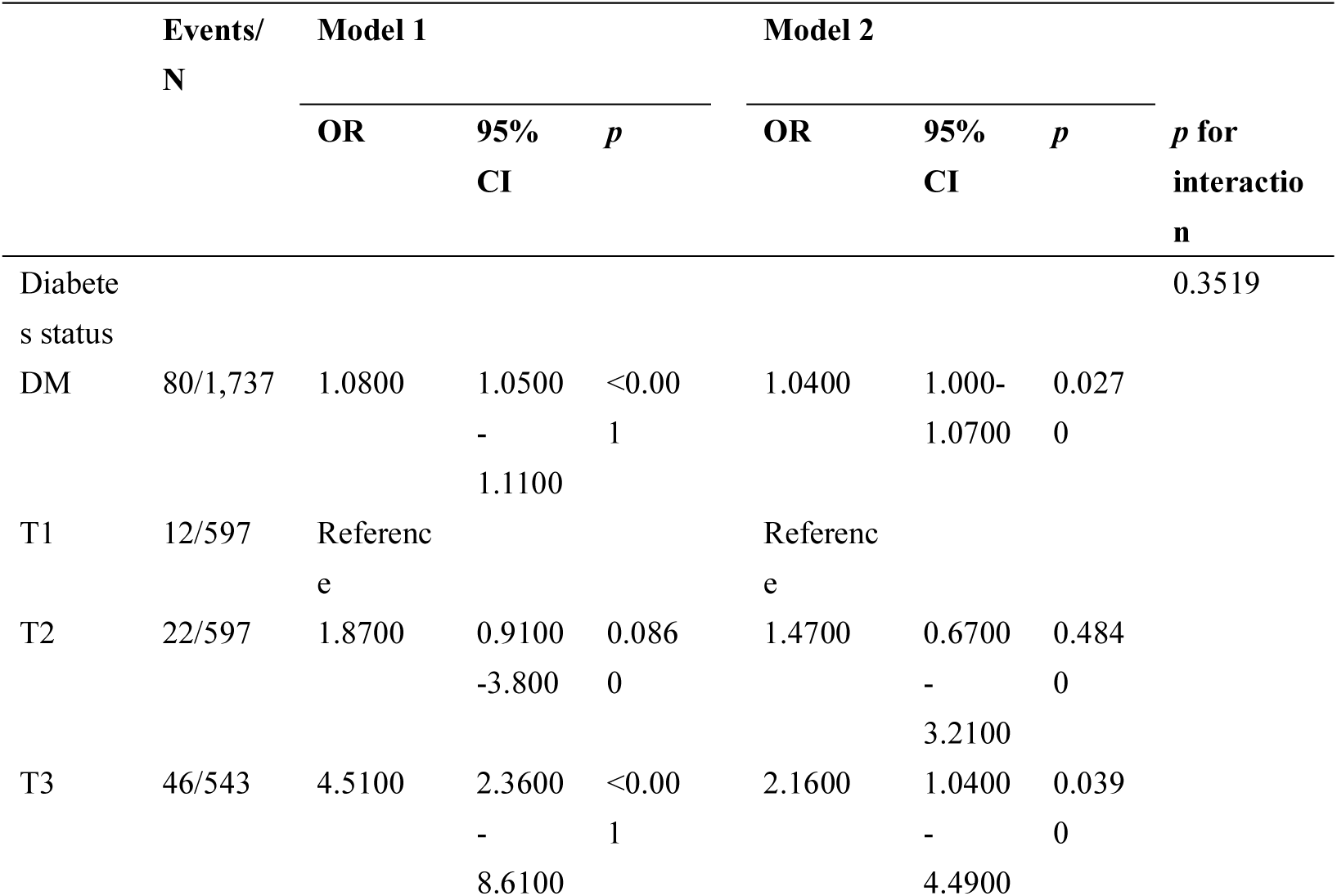
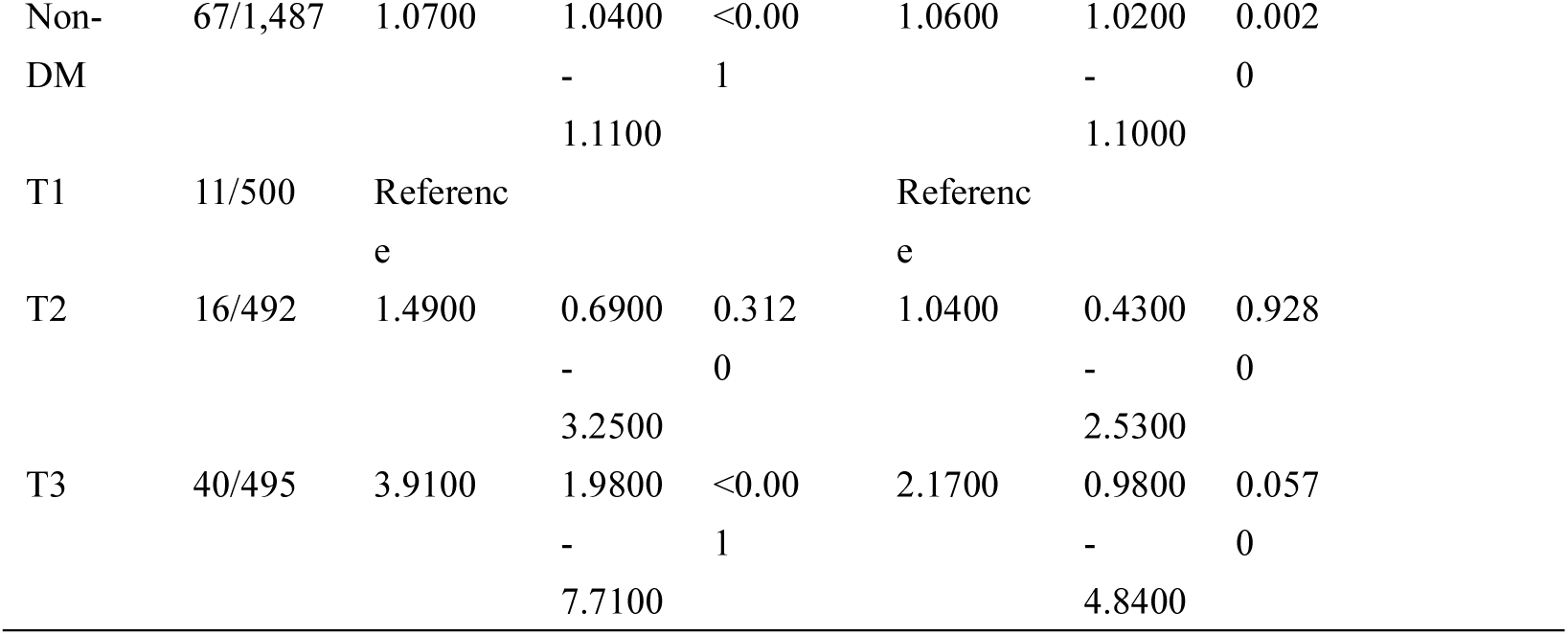
Relationship between SYNTAX score and in-hospital cardiac arrest incidence in DM and non-DM patients.

Table 7 displays the relationship between SS and IHCA incidence in elderly (≥60 years) and non-elderly (<60 years) patients. No interaction was found between SS and IHCA incidence in these subgroups (*p* = 0.5054). Among elderly patients, both Model 1 (OR = 1.0700; 95% CI = 1.0400–1.0900; *p* < 0.001) and Model 2 (OR = 1.0300; 95% CI = 1.0100–1.0700; *p* = 0.0210) showed a significant association between SS and IHCA incidence. Among non-elderly patients, both Model 1 (OR = 1.0900; 95% CI = 1.0500– 1.1300; *p* < 0.001) and Model 2 (OR = 1.0500; 95% CI = 1.000–1.1000; *p* = 0.0470) demonstrated a significant association.

**Table 7.**
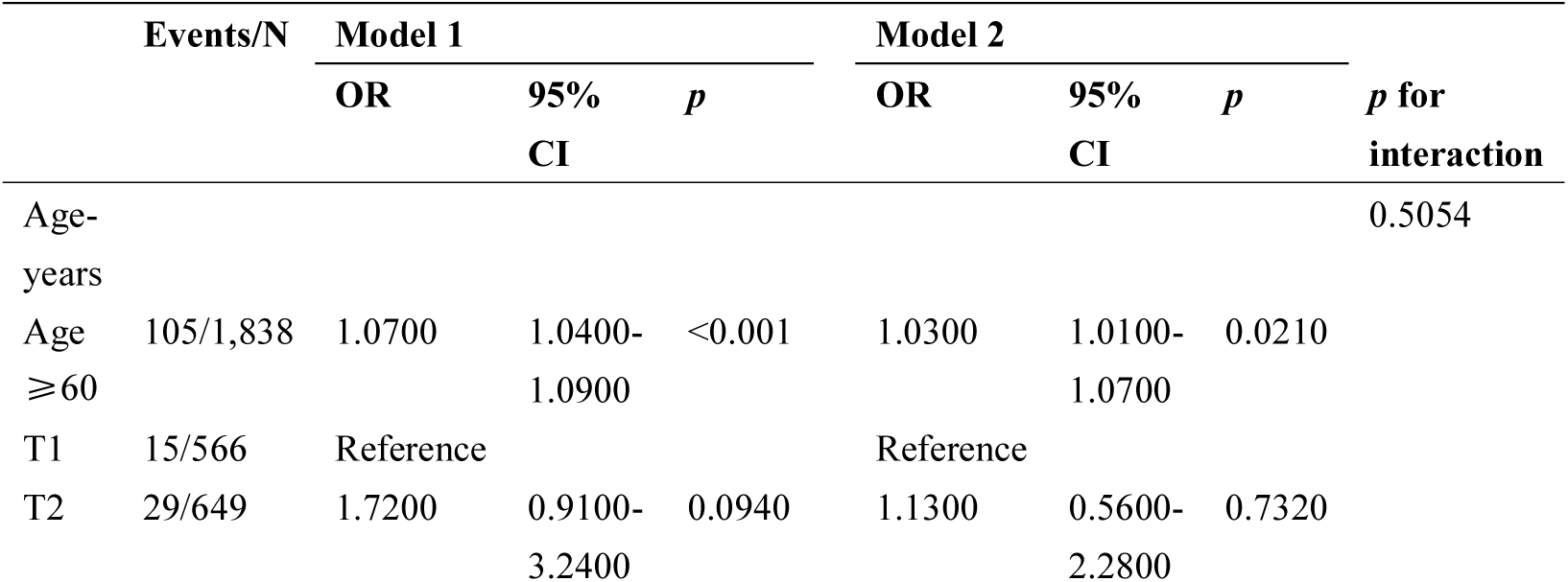
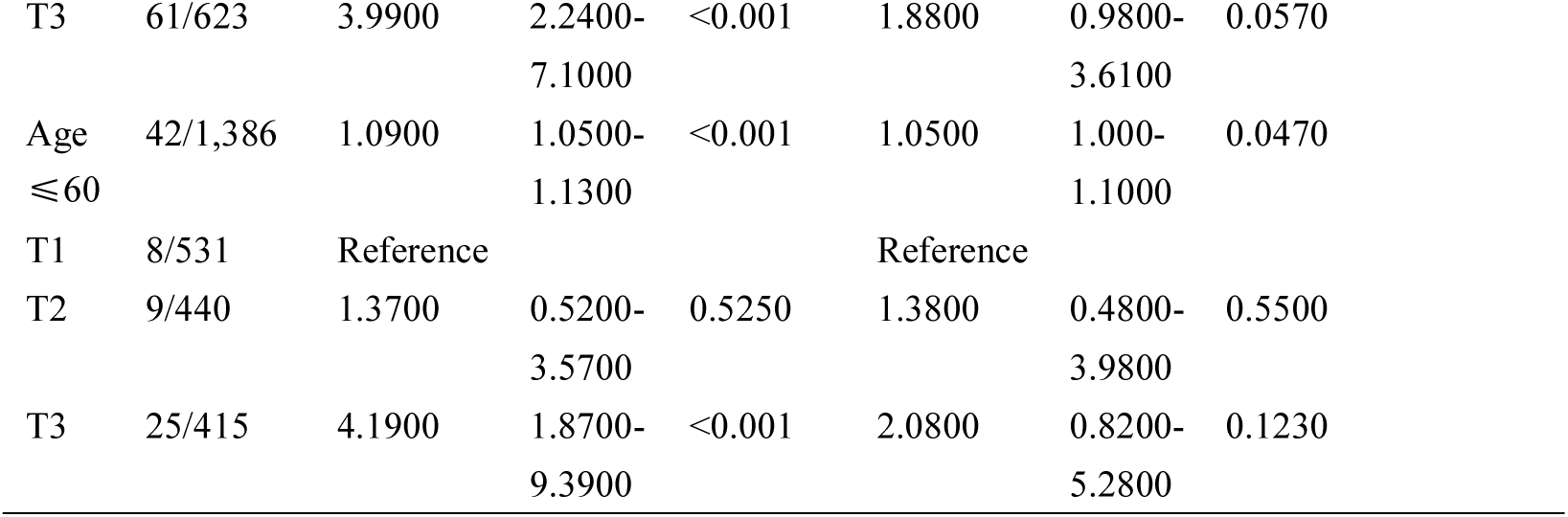
Relationship SYNTAX score and in-hospital cardiac arrest incidence in Age≥60 and Age ≤60 patients.

|  | Events/N | Model 1 |  |  | Model 2 |  |  | *p* for interaction |
| --- | --- | --- | --- | --- | --- | --- | --- | --- |
|  |  | OR | 95% CI | *p* | OR | 95% CI | *p* |  |
| Age-<br>years |  |  |  |  |  |  |  | 0.5054 |
| Age $\geq 60$ | 105/1,838 | 1.0700 | 1.0400–1.0900 | <0.001 | 1.0300 | 1.0100–1.0700 | 0.0210 | |
| T1 | 15/566 | Reference |  |  | Reference |  |  |  |
| T2 | 29/649 | 1.7200 | 0.9100–3.2400 | 0.0940 | 1.1300 | 0.5600–2.2800 | 0.7320 |  |

|  |  |  |  |  |  |  |  |
| --- | --- | --- | --- | --- | --- | --- | --- |
| T3 | 61/623 | 3.9900 | 2.2400-7.1000 | <0.001 | 1.8800 | 0.9800-3.6100 | 0.0570 |
| Age<br>≤60 | 42/1,386 | 1.0900 | 1.0500-1.1300 | <0.001 | 1.0500 | 1.000-1.1000 | 0.0470 |
| T1 | 8/531 | Reference |  |  | Reference |  |  |
| T2 | 9/440 | 1.3700 | 0.5200-3.5700 | 0.5250 | 1.3800 | 0.4800-3.9800 | 0.5500 |
| T3 | 25/415 | 4.1900 | 1.8700-9.3900 | <0.001 | 2.0800 | 0.8200-5.2800 | 0.1230 |

Table 8 illustrates the relationship between SS and IHCA incidence in male and female patients. No interaction was observed between SS and IHCA incidence across gender subgroups (*p*=0.8538). Among male patients, Model 1 showed that SS was significantly associated with IHCA risk (OR = 1.0700; 95% CI = 1.0500–1.0900; *p* < 0.001), with the T3 group having 4.06 times the incidence of the T1 group (OR = 4.0600; 95% CI = 2.3800–6.9200; *p* < 0.001). Model 2 also indicated a significant association (OR = 1.0500; 95% CI = 1.0200–1.0700; *p* = 0.0020), with the T3 group having 2.15 times the incidence of the T1 group (OR = 2.1500; 95% CI = 1.1800–3.9300; *p* = 0.0130).

**Table 8.**
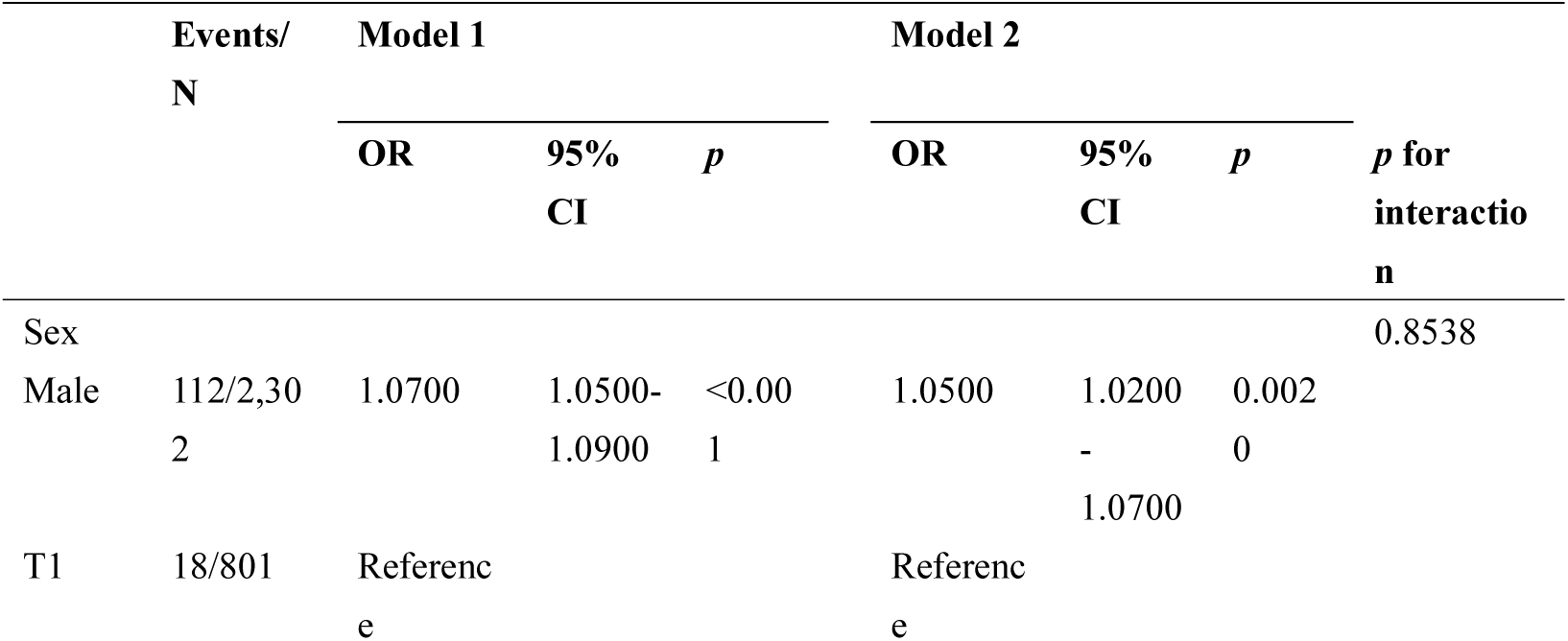
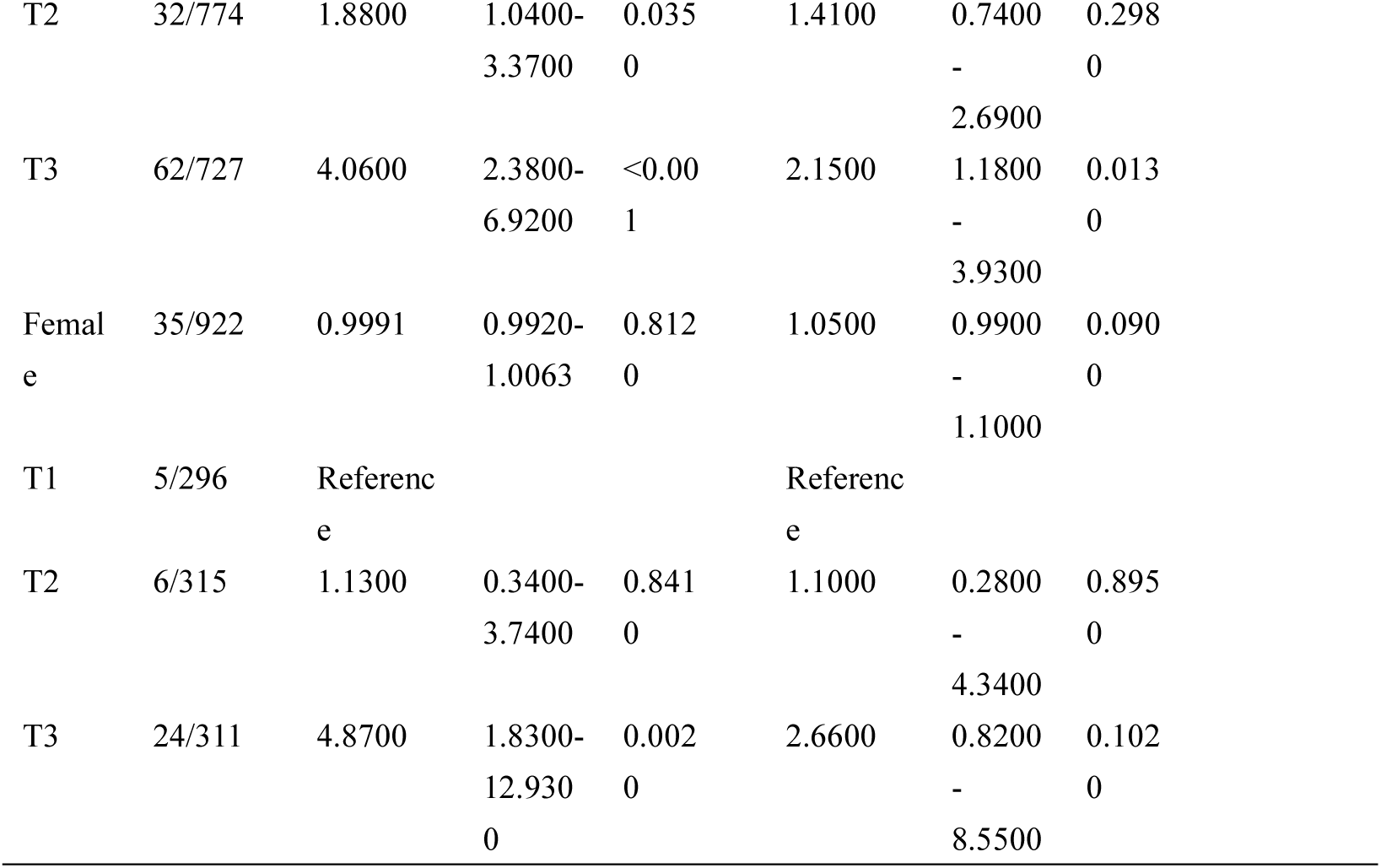
Relationship between SYNTAX score and in-hospital cardiac arrest incidence in Male and Female patients.

|  | Events/<br>N | Model 1 |  |  | Model 2 |  |  | *p* for<br>interaction |
| --- | --- | --- | --- | --- | --- | --- | --- | --- |
|  |  | OR | 95%<br>CI | *p* | OR | 95%<br>CI | *p* |  |
| Sex |  |  |  |  |  |  |  | 0.8538 |
| Male | 112/2,302 | 1.0700 | 1.0500-1.0900 | <0.001 | 1.0500 | 1.0200<br>-<br>1.0700 | 0.0020 |  |
| T1 | 18/801 | Reference |  |  | Reference |  |  |  |

|  |  |  |  |  |  |  |  |
| --- | --- | --- | --- | --- | --- | --- | --- |
| T2 | 32/774 | 1.8800 | 1.0400-3.3700 | 0.035<br>0 | 1.4100 | 0.7400<br>-<br>2.6900 | 0.298<br>0 |
| T3 | 62/727 | 4.0600 | 2.3800-6.9200 | <0.00<br>1 | 2.1500 | 1.1800<br>-<br>3.9300 | 0.013<br>0 |
| Femal<br>e | 35/922 | 0.9991 | 0.9920-1.0063 | 0.812<br>0 | 1.0500 | 0.9900<br>-<br>1.1000 | 0.090<br>0 |
| T1 | 5/296 | Referenc<br>e |  |  | Referenc<br>e |  |  |
| T2 | 6/315 | 1.1300 | 0.3400-3.7400 | 0.841<br>0 | 1.1000 | 0.2800<br>-<br>4.3400 | 0.895<br>0 |
| T3 | 24/311 | 4.8700 | 1.8300-12.930<br>0 | 0.002<br>0 | 2.6600 | 0.8200<br>-<br>8.5500 | 0.102<br>0 |

### The Incremental Effect of SS on Predicting the Incidence of IHCA

ROC curves were constructed to evaluate the baseline risk model (including Age, DBP, Stent Diameter, eGFR, ABG, hs-CRP, HbA1c, LVEF, Male, Smoking, Previous MI, LCX disease, RCA disease, Bifurcation lesion, CTO disease, Thrombolytic therapy, Hypertension, Dyslipidemia, Previous stroke, Insulin, Oral hypoglycemic drugs, Aspirin, and P2Y12 inhibitors) and the predictive ability of the baseline model combined with HbA1c, ABG, and SS for IHCA incidence in ACS patients undergoing PCI (Figure 3). Table 9 lists the C-statistic, NRI, and IDI. The results indicate that SS provides significant incremental predictive value to the baseline risk model (NRI: 0.3537 [0.1906–0.5167], *p* < 0.001; IDI: 0.0104 [0.0025–0.0183], *p* = 0.0097).

**Figure 3.**
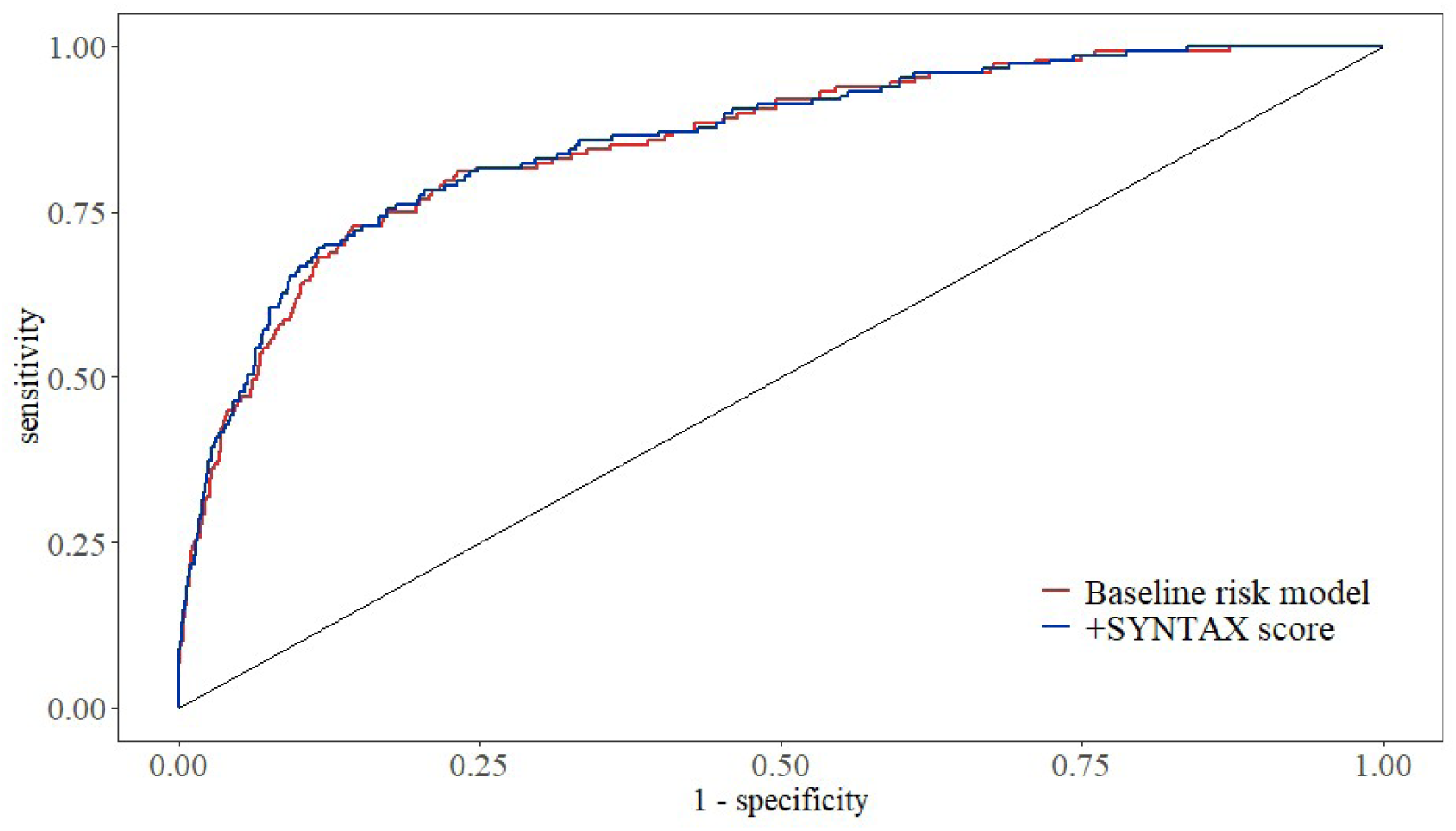
Receiver operating characteristic curves assessing the predictive ability of SYNTAX score for incidence of in-hospital cardiac arrest. Baseline risk model vs + SYNTAX score in ACS patients treated with PCI.

**Table 9.** Incremental predictive value and predictive power of various models with NRI, IDI, and C-statistics.

**C-statistics**
| Model | C-statistic | $p$ | NRI | $p$ | IDI | $p$ |
| --- | --- | --- | --- | --- | --- | --- |

|  | (95% CI) |  | (95% CI) |  | (95% CI) |  |
| --- | --- | --- | --- | --- | --- | --- |
| Baseline risk model | 0.8543<br>(0.8218-0.8867) | Ref | Ref |  | Ref |  |
| +SYNTAX score | 0.8581<br>(0.8256-0.8906) | 0.3540 | 0.3537<br>(0.1906-0.5167) | <0.001 | 0.0104<br>(0.0025-0.0183) | 0.0097 |

## Discussion

In this study, the SS was used to quantitatively assess the severity of coronary artery lesions in patients with ACS undergoing PCI. The results indicated that SS was significantly associated with the incidence of in-hospital cardiac arrest (IHCA). RCS analysis revealed a dose-response relationship between SS and IHCA incidence. Furthermore, incorporating SS into the baseline risk model provided incremental predictive value for IHCA in this patient population.

The potential mechanisms by which elevated SS increases IHCA risk involve several interrelated pathophysiological pathways. First, SS comprehensively evaluates lesion characteristics, including location, number, severity, chronic total occlusion (CTO), bifurcation, calcification, and diffuse disease; a higher score typically corresponds to more extensive myocardial ischemia, which can increase transmurality repolarization dispersion and generate injury currents, thereby triggering ventricular tachycardia or fibrillation^21,22^. Previous studies have confirmed that CTO and high SS predict mortality following cardiac arrest^23^, consistent with the findings of this study. Second, SS is significantly associated with markers of systemic inflammation, such as the neutrophil-to-lymphocyte ratio, systemic immune-inflammatory index, Naples score, and triglyceride-glucose index^24–27^. Hyperinflammation promotes plaque rupture, platelet activation, and microvascular dysfunction. Given that ACS is characterized by acute plaque instability, the diffuse lesions present in patients with high SS make untreated plaques more prone to acute rupture during hospitalization, triggering ischemic malignant arrhythmias. Furthermore, PCI procedures in patients with high SS are typically more complex. Interventions such as multiple stent placements, bifurcation management, and CTO recanalization increase procedure duration and operational risks^28^. These factors are associated with a higher incidence of post-procedural slow flow or no-reflow^29^, leading to acute myocardial injury. Previous studies have also suggested an increased risk of post-PCI bleeding and in-stent thrombosis^28,30^. Additionally, achieving complete revascularization is often difficult in patients with high SS; a residual SS > 8 is significantly associated with 5-year mortality^31–33^. Pathologies left untreated or inadequately treated during hospitalization can lead to acute ischemia under conditions of stress, inflammation, and inadequate anticoagulation, contributing significantly to IHCA. Finally, in cases of acute coronary occlusion, excessive sympathetic nervous system activation has a clear arrhythmogenic effect, whereas vagal nerve activation exerts an anti-fibrillation effect^21^. A more extensive area of ischemia may exacerbate autonomic imbalance, promoting the progression of ventricular tachycardia to ventricular fibrillation^22^. These mechanisms are not independent but act in concert through a continuous pathway: “high anatomical complexity → extensive myocardial ischemia/necrosis + inflammatory activation + incomplete revascularization + perioperative ischemic events + autonomic imbalance → myocardial electrical instability and/or deterioration of mechanical function → IHCA.” Multiple studies have confirmed that SS is a robust predictor of short-term and long-term mortality in patients with STEMI ^34–38^. Building on this foundation, this study focuses on IHCA—a more time-sensitive and mechanistically direct adverse event—thereby expanding the prognostic applications of SS. A subgroup analysis of the ACUITY trial showed that SS is an independent predictor of 1-year mortality and myocardial infarction in patients with NSTEMI^39^. This study further limited the risk prediction window to the hospitalization period, suggesting that the risk stratification value of this scoring system is fully demonstrated during the acute phase. In patients with severe cardiogenic shock, the CULPRIT-SHOCK trial confirmed that SS strongly predicts 30-day mortality or the need for renal replacement therapy, as well as 1-year mortality, in patients with infarction-related cardiogenic shock^40^. Although this study included patients across a broader spectrum of severity, the results were consistent with those of the aforementioned studies, indicating that the prognostic value of SS remains robust across different severity levels. This study offers unique and complementary value compared to previous research on cardiac arrest and sudden cardiac death. Demirkıran et al. found that SS was significantly higher in patients who died after cardiac arrest than in survivors (16.3 ± 3.8 vs. 13.6 ± 1.9), and that CTO could predict mortality following cardiac arrest secondary to STEMI^23^. In contrast, this study advanced the assessment time point to admission and the PCI hospital stay, finding that SS also predicts IHCA, thereby forming a complementary chain of evidence. Yannopoulos et al. observed that 84% of patients with refractory out-of-hospital ventricular fibrillation/ventricular tachycardia-related cardiac arrest had significant coronary artery disease, with a mean SS as high as 29.4^41^. Franco et al. further found that SS in patients with out-of-hospital cardiac arrest increased with resuscitation difficulty, rising from 10.2 in conscious patients to 26.8 in those requiring extracorporeal membrane oxygenation (ECMO) support^42^. Unlike these studies of out-of-hospital scenarios, this study focuses on IHCA, which has been relatively understudied, and suggests that such events are highly dependent on coronary anatomical burden. In a 20-year follow-up study of patients with early-onset myocardial infarction, Bricoli et al. found that SS is an independent predictor of sudden cardiac death^43^. This study extends the predictive logic for long-term sudden cardiac death to episodes of acute cardiac arrest during hospitalization, supporting the existence of a continuous chain of events: “high anatomical complexity → acute electrical instability → malignant arrhythmias.” Having established an independent association between SS and IHCA, it is important to examine their dose-response relationships and risk thresholds.

The RCS analysis in this study showed that SS was associated with IHCA risk in an approximately linear manner (nonlinearity *p* = 0.906), with an inflection point of approximately 23.975. This value is close to the medium-risk/high-risk cutoff values of 22 or 23 widely used in previous studies^37,38,44^. This consistency suggests that an SS value in the range of 22 to 24 may correspond to a common biological threshold at which coronary anatomical burden triggers clinically adverse events. When SS < 23.975, each increase of 1 standard deviation was associated with a 30% increase in the risk of IHCA (OR = 1.30, 95% CI: 1.03–1.63). When the score was greater than 23.975, for every additional standard deviation, the OR decreased to 0.999 (95% CI: 0.625–1.510). This phenomenon may be related to the small sample size in the high-risk subgroup, the low number of events, and early survival bias among extremely high-risk patients; it requires validation in future studies with larger sample sizes. In the subgroup analysis, all *p*-values for interactions were greater than 0.05; the differing results may be due to insufficient sample size. Based on the above evidence, SS, as an angiographic quantification tool that can be rapidly calculated during routine PCI procedures, has clear clinical applicability and does not incur additional testing costs. Guidelines in Europe and the United States have already recommended its use in decision-making regarding revascularization strategies^38,45,46^. This study further expands its application to the risk stratification of in-hospital complications. Specifically, patients with an SS > 23.975 or in the top third (>18) have a significantly increased risk of IHCA. It is recommended that such patients be transferred to the coronary care unit (CCU) or intensive care unit (ICU) after PCI for continuous ECG monitoring, along with enhanced monitoring of electrolyte levels, acid-base balance, and hemodynamics. For patients with high-risk characteristics such as DM, a history of myocardial infarction, CTO, or multivessel disease, SS can further refine risk stratification to guide the selection of perioperative anticoagulation strategies, heart rate and blood pressure management, and early mechanical circulatory support when necessary. When conditions permit, the possibility of complete revascularization or consultation with a cardiac surgeon should be evaluated to minimize the risk of residual ischemia-related cardiac arrest^47,48^. Consistent with previous approaches to developing derived scores^49–52^, this study supports the integration of anatomical scores with clinical variables to construct a multidimensional comprehensive predictive model. It is important to emphasize that SS should not be used in isolation; rather, a comprehensive assessment must be made based on ACS type, hemodynamic status, biochemical markers, and Holter monitoring results. Additionally, this scoring method is subject to inter-observer variability; in critical decision-making scenarios, it is recommended that a central laboratory conduct a review. In the future, larger-scale prospective cohort studies should be conducted to investigate the predictive value of SS for cardiovascular outcomes in patients with ACS.

### Strengths and limitations

To our knowledge, this is the first study to use RCS analysis to identify a dose-response relationship between SS and IHCA in patients with ACS and to evaluate the linear correlation between SS and IHCA events. However, this study has several limitations. As a single-center retrospective cohort study, selection bias and information bias are difficult to completely avoid. The center’s PCI experience, postoperative monitoring protocols, and patient recruitment may limit the generalizability of the results. The retrospective design also means that residual confounding and reverse causality cannot be completely ruled out. Although the association between SS and IHCA remains robust after adjusting for multiple factors, a causal relationship cannot be directly inferred. SS calculations rely on the quality of imaging during procedures and observer interpretation, and lack validation of consistency across core laboratories. Scores may change following PCI; this study did not evaluate the dynamic effects of residual SS and therefore could not capture the contribution of residual ischemia to IHCA after revascularization. In addition, the study lacked data on postoperative dynamic hemodynamic parameters and serial echocardiographic changes; these omitted variables may have a residual confounding effect on the risk of IHCA.

## Conclusions

In ACS patients undergoing PCI, SS is significantly associated with the incidence of IHCA and may serve as an effective predictor. Furthermore, incorporating SS into baseline risk models provides incremental predictive value for IHCA. Future large-scale, multicenter, prospective studies are warranted to validate the predictive utility of SS, while the mechanisms underlying the observed dose-response relationship require further investigation.

## List of Abbreviations

SYNTAX score, SS; In-hospital cardiac arrest, IHCA; Diabetes Mellitus, DM; Percutaneous Coronary Intervention, PCI; ST-Elevation Myocardial Infarction, STEMI; Non-ST-Elevation Myocardial Infarction, NSTEMI; Unstable Angina, UA; Triglycerides, TG; Admission Blood Glucose, ABG; Haemoglobin A1c, HbA1c; Estimated Glomerular Filtration Rate, eGFR; Acute Coronary Syndrome, ACS; Creatinine, Cr; Odds Ratio, OR; Confidence Interval, CI; Receiver Operating Characteristic, ROC; Area Under the Curve, AUC; Net Reclassification Improvement, NRI; Integrated Discriminant Improvement, IDI; Myocardial Infarction, MI;

## Declarations

### Ethics approval and consent to participate

The study was conducted in accordance with the Declaration of Helsinki and was approved by the Ethics Committee of the First Affiliated Hospital of Jinan University.

### Consent for publication

Not applicable.

### Availability of data and materials

Due to privacy and ethical constraints, the datasets generated and analysed in this study are not publicly available but can be obtained from the corresponding author.

### Authors’ contributions

KL designed the study, analysed the data, and wrote the manuscript; HXC collected and interpreted the data; HWY and JG critically revised the manuscript for important intellectual content. All authors read and approved the final manuscript.

## Acknowledgements

We thank all the members who contributed to this study.

